# Prediction and predictor elucidation of the onset of metabolic syndrome among young workers using machine learning techniques: A nationwide study in Japan

**DOI:** 10.1101/2021.10.21.21265259

**Authors:** Miyuki Suda, Tadao Ooka, Zentaro Yamagata

## Abstract

**Objectives:** Predictive models for the onset of metabolic syndrome (MS) among people in their 30s are scarce. This study aimed to construct a highly accurate model to predict MS onset by age 40 years and to identify the important predictors of MS onset using health checkup data of Japanese company employees aged 30 and 35.

**Methods:** The study included 6,048 Japanese employees aged 40 years who had undergone periodic health examinations over 10 years. We developed prediction models for MS onset using machine learning methods including the random forest and logistic regression, and evaluated the models using the receiver operating characteristics and precision-recall curves. For the random forest models, the variable importance of each explanatory variable was calculated to identify important predictors of MS onset.

**Results:** The random forest had higher predictive power than logistic regression in all models, although the differences were non-significant. Regarding important predictors, diastolic blood pressure was the most important predictor of MS onset for men aged 30 and 35 years, while body mass index was the most important predictor for women aged 30 and 35 years.

**Conclusions:** We created a machine learning model to predict MS onset at age 40 with high accuracy from health examination data obtained at ages 30 or 35. Sex differences in important predictors of MS onset was shown by the variable importance indices of the random forest. Applying our model in routine healthcare management should provide early and appropriate health interventions to prevent MS onset in young people.

## 1. INTRODUCTION

Metabolic syndrome (MS) is a combination of metabolic disorders, including obesity, hyperglycemia, hypertension, and lipid abnormalities, which predispose to diabetes and cardiovascular disease.^1^ There are over 1 billion persons with MS worldwide, indicating that a considerable proportion of the world’s population is at risk of MS. ^2^Therefore, preventing MS is an important global issue from a public health perspective.

In 1999, the World Health Organization (WHO) proposed criteria for the diagnosis of MS.^3^Since then, two approaches to the diagnostic criteria for MS have been advanced. The first is based on the WHO concept, which includes insulin resistance and visceral fat.^3,4^ The second is based on the overlap of risk factors for cardiovascular disease, including obesity and hypertension.^5,6^ The former concept is mainly used in Japan^4^, while the latter is the mainstream in the U.S. and in European countries.

In Japan, people aged 40 years or older have the option to receive health guidance as a preventive measure against MS in accordance with national law. However, there are no MS prevention measures for people aged less than 40 years. Previous studies have reported that the basis for MS is established before the age of 40^7^ and that the lifestyle in the 30s is associated with the development of MS in the 40s. ^8-10^ In addition, a previous study on weight control suggests that health education by age 35 leads to weight reduction at age 40.^11^ Furthermore, other studies have reported the importance of developing a good lifestyle habits before the age of 40 to prevent weight gain in older age.^12^

Therefore, identifying individuals at high risk of MS in their 30s and intervening to improve their lifestyle habits should be useful in preventing MS. However, most previous studies for identifying MS risk were conducted with the data of people aged over 40 years because of the difficulty of obtaining health information from young people. Moreover, to the best of our knowledge, there is no report on the construction of predictive models for MS onset in young people in their 30s although this seems important for detecting early MS development.

In this study, we obtained longitudinal health examination data of men and women from a Japanese company; the data collection began in their 30s and had been recorded for over 10 years. We then built a prediction model for MS onset for young people by applying machine learning methods to these data. In addition, by using a highly interpretable machine learning method ^13^and comparing the detected variables to clinically well-known factors, we confirmed the validity of the models and identified important factors associated with the development of MS in men and women in their 30s.

## 2. METHODS

### 2.1. Study design and participants

The study included Japanese employees who were 30 years old in 2008 or 2009 and underwent continuous periodic health examinations conducted from 2008 to 2019 by *Health Insurance Association A*, which is in charge of health management among 525 business sites throughout the nation (**Figure 1**).

**Figure 1.**
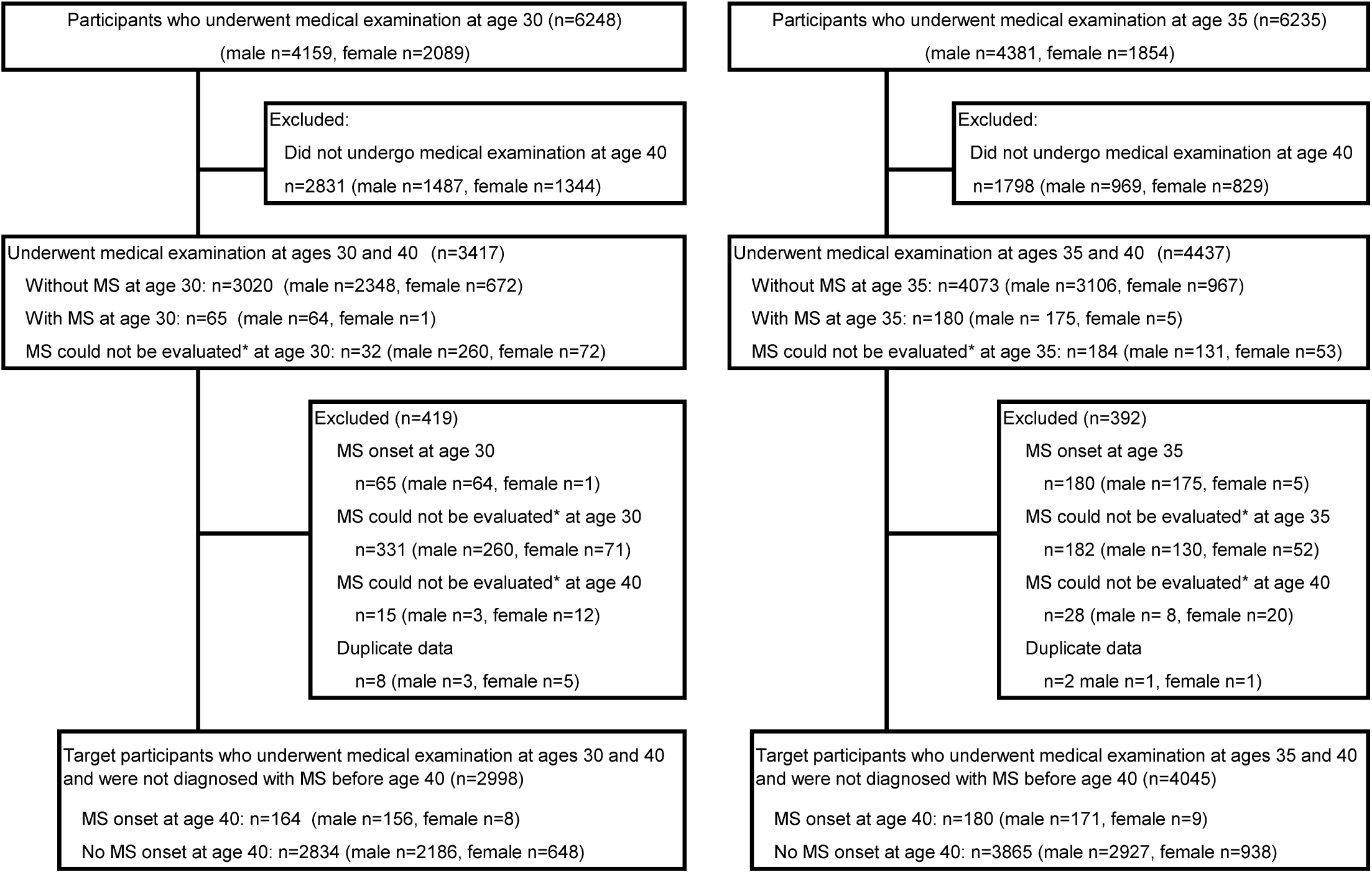
Participant selection flow ^*^Study participants who could not be assessed for MS because of some missing values. MS, metabolic syndrome

For each year, we excluded participants who could not be evaluated for MS because of missing values or who were already identified with MS at age <40. If participants underwent two medical examinations in the same year, only the data point with fewer missing values or the first data of the year (when the number of missing values was the same in the two examinations) were included in the analysis.

Finally, we prepared two datasets for the analyses in this study. The first dataset was created by combining the data from the health examination at age 30 with the MS evaluation data at age 40. The second dataset was created by combining data from health examinations at age 35 with the MS evaluation data at age 40. We used these two datasets to construct prediction models for MS onset at age 40 and to examine important predictors of MS onset.

### 2.2. Measures

#### 2.2.1. Outcome

The main outcome was the onset of MS at age 40. The diagnostic criteria for MS were based on the 2005 Japanese Journal of Internal Medicine criteria,^4^ which is the most widely used criteria in Japan. For the diagnosis of MS, a notably large abdominal circumference (male ≥85 cm, female ≥90 cm) was set as a mandatory item, and at least two items among blood pressure (systolic blood pressure ≥ 130 mmHg or diastolic blood pressure ≥ 85 mmHg), lipid levels (triglycerides ≥ 150 mg/dL or high-density lipoprotein cholesterol [HDL-C] ≤ 40 mg/dL), and blood glucose level (fasting blood glucose ≥ 110 mg/dL) needed to exceed the standard values. In addition, patients receiving medication for blood pressure, lipids, and blood glucose were considered to meet the criteria for each item, even if the standard values were not exceeded.

#### 2.2.2. Predictive variables

In constructing the prediction model, we used 16 examination items or 12 interview items from the health examination prescribed by Japanese law. The examination items included (1) body mass index; (2) waist circumference; (3) systolic blood pressure; (4) diastolic blood pressure; (5) high-density lipoprotein cholesterol (HDL-C); (6) low-density lipoprotein cholesterol (LDL-C); (7) triglycerides; (8) alanine aminotransferase (ALT); (9) aspartate aminotransferase (AST); (10) γ-glutamyl transpeptidase (γ-GTP); (11) blood glucose; (12) hematocrit; (13) hemoglobin; (14) red blood cells; (15) white blood cells; and (16) uric acid.

The interview items included (1) daily alcohol consumption; (2) having breakfast; (3) paying attention to nutritional balance; (4) walking for more than 1 h per day; (5) walking speed; (6) intention to improve health; (7) eating before bed; (8) restful sleep; (9) eating too fast; (10) cigarette smoking; (11) weight gain >10 kg; and (12) exercising more than twice a week

Physical measurements and blood tests were treated as continuous variables, while all questionnaire items were treated as binary variables.

#### 2.2.3. Statistical analysis

The prediction models for MS onset were created using machine learning methods, including random forest (RF) and logistic regression (LR), and were evaluated with the area under the receiver operating characteristics curve (AU-ROC) and the area under the precision recall curve (AU-PRC).

Training data were used to create the machine learning models, and test data were used to check model accuracy. The training and test datasets were randomly selected from the original dataset in a 4:1 for men and 1:1 ratio for women because of the small number of MS cases for women. In all models, the outcome was the presence or absence of MS at age 40, and the 28 items, including all examination items and interview items, were used as explanatory variables.

In the construction of the prediction model, RF modelling was performed using the *randomForest* package of the statistical software R. The number of decision trees (*ntree* parameter) was set to 1000, and the number of features used to create the decision trees (*mtry* parameter) was automatically set by the R *caret* package.^14^ The analysis was performed with 10-segment cross-validation.

In the LR model, in order to compare the performance under the same conditions as RF, all variables were used as explanatory variables with the forced entry method without selecting variables based on prior knowledge. To avoid the problem of complete separation caused by the small number of female cases, Firth’s bias-reduced logistic regression was used to create the LR model for the 30-year-old women. ^15^

When we created RF models, the variable importance of each explanatory variable was calculated to identify important predictors of MS. In the calculation of variable importance, we used RF with conditional inference trees (using the cForest package ^16^) because RF has a tendency to underestimate categorical variables with fewer categories. In addition to creating the predictive models, we used multidimensional scaling (MDS) to evaluate the similarity between MS and non-MS patients. ^17^ All analyses were performed using R version 3.6.1 (R Foundation for Statistical Computing, Vienna, Austria), and set the significance level to 0.05.

#### 2.2.4. Ethical Considerations

In this study, we used health checkup data collected by Health Insurance Association A. All datasets used in this study were anonymized and statistically processed in systems that were unconnected to any external networks or the Internet. Personal information was strictly protected and managed in accordance with the ethical guidelines established by the government (Ethical Guidelines for Medical Research Involving Human Subjects).^18^ This study was approved by the Ethics Committee of the University of Yamanashi (Ethics Committee receipt number R01688). The study was also approved by the Ethics Committee of Health Insurance Association A (receipt number 2019-002). All participants were provided the opportunity to opt out.

## 3. RESULTS

Of the 6248 participants who underwent a physical examination at age 30 and 6235 participants who underwent a physical examination at age 35, 2998 (2342 men and 656 women) at age 30 and 4045 (3098 men and 947 women) at age 35 had MS assessment data at age 40 without developing MS (**Figure 1**).

When the blood test data at age 30 were compared between the MS-onset and non-MS-onset groups at age 40, all the blood test data except HDL-C were significantly higher in the MS-onset group, and HDL-C was significantly lower in the MS-onset group (**Table 1**). The blood test data at age 35 were also compared between the MS-onset and non-MS-onset groups at age 40, with the same results as at age 30.

**Table 1.**
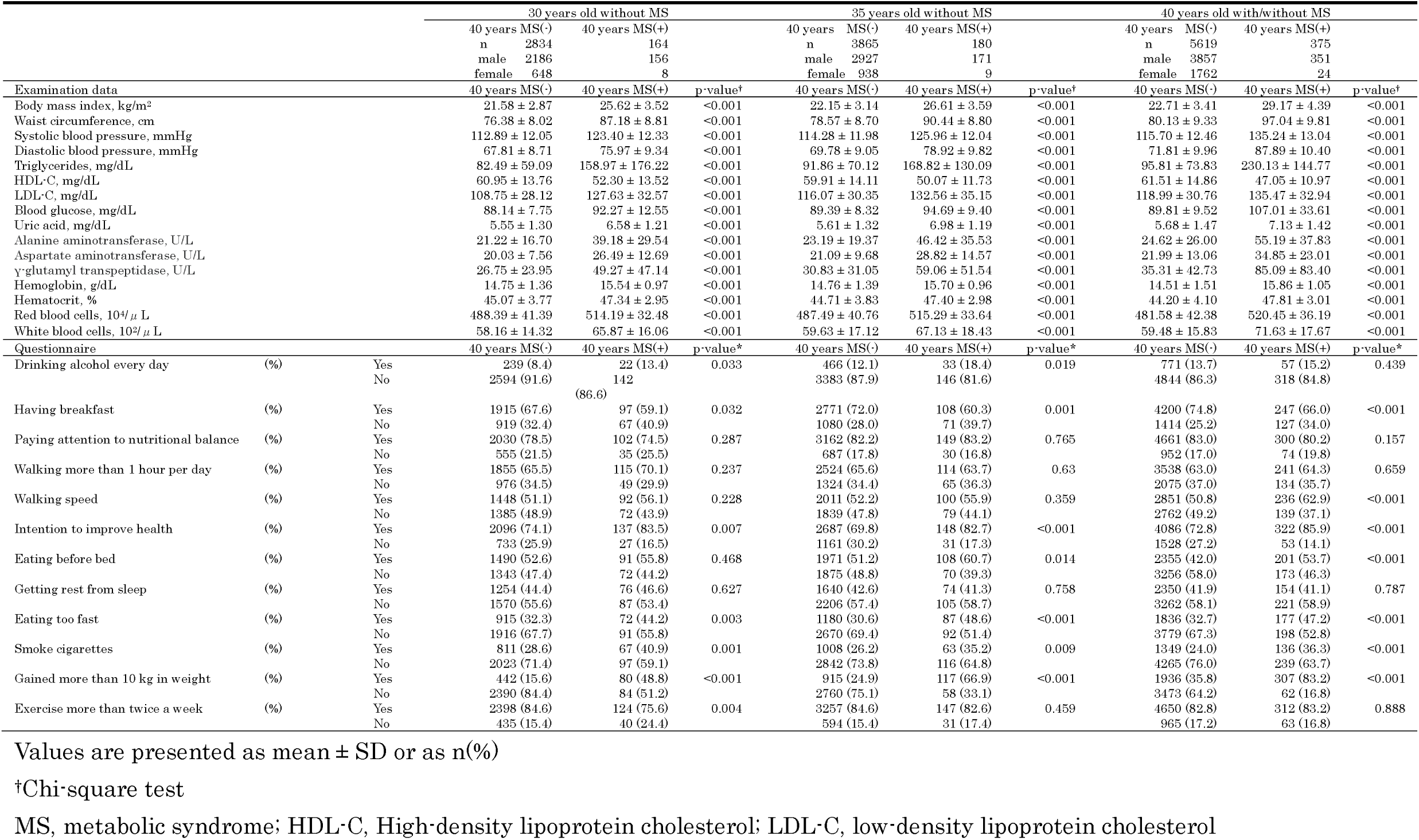
Characteristics of study participants.

In terms of the health examination questions at ages 30 and 35, we had significantly higher prevalence of “yes” among 6questions (“Drinking alcohol every day” “Not having breakfast,” “Intention to improve,” “Gained more than 10 kg in weight,” “Eating too fast,” and “Smoke cigarettes”) (**Table 1**). We compared the predictive accuracy of the models using AU-ROC and AU-PRC, and we found that the RF had a higher AU-ROC and AU-PRC than LR in all the models, although the differences in accuracy were not significant (**Table 2 and Figure 2**).

**Table 2.**
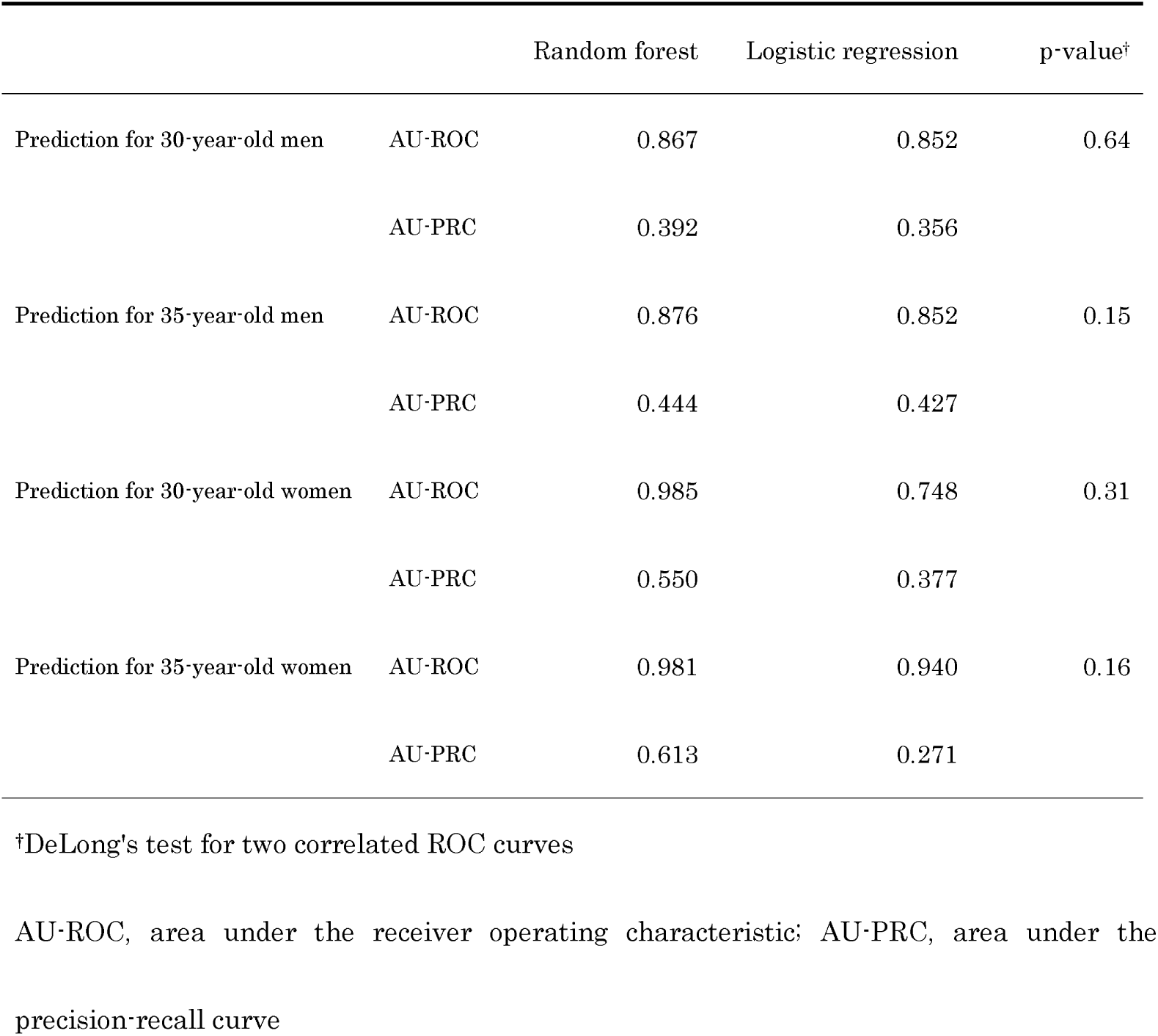
Predictive accuracy of the random forest and logistic regression models.

|  |  | Random forest | Logistic regression | p-value<sup>†</sup> |
| --- | --- | --- | --- | --- |
| Prediction for 30-year-old men | AU-ROC | 0.867 | 0.852 | 0.64 |
|  | AU-PRC | 0.392 | 0.356 |  |
| Prediction for 35-year-old men | AU-ROC | 0.876 | 0.852 | 0.15 |
|  | AU-PRC | 0.444 | 0.427 |  |
| Prediction for 30-year-old women | AU-ROC | 0.985 | 0.748 | 0.31 |
|  | AU-PRC | 0.550 | 0.377 |  |
| Prediction for 35-year-old women | AU-ROC | 0.981 | 0.940 | 0.16 |
|  | AU-PRC | 0.613 | 0.271 |  |

<sup>†</sup>DeLong's test for two correlated ROC curves
AU-ROC, area under the receiver operating characteristic; AU-PRC, area under the precision-recall curve

**Figure 2.**
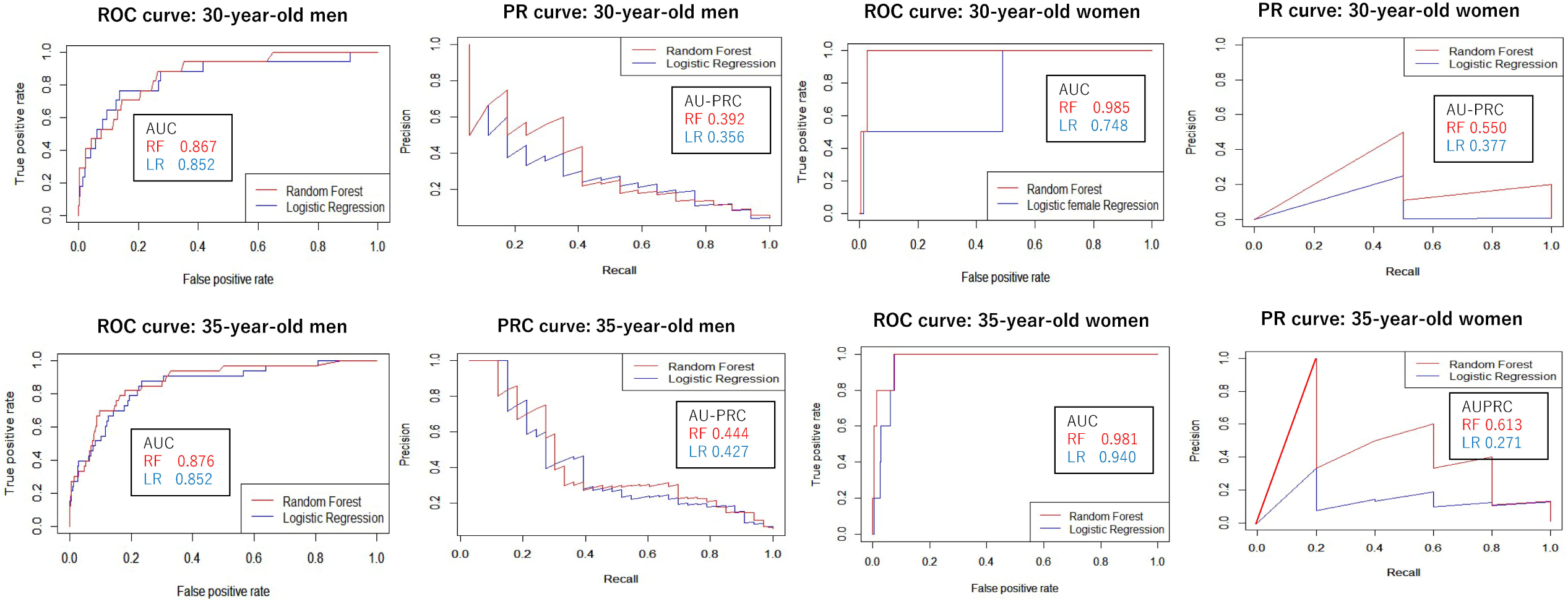
Receiver operating characteristic (ROC) curve and precision-recall (PR) curve for predicting the onset of metabolic syndrome via the random forest and logistic regression models.

We assessed the importance of the predictors by calculating the variable importance of the explanatory variables in the RF models, and diastolic blood pressure was shown to be the most important predictor of MS onset in men aged 30 and 35 years. LDL-C, BMI, HDL-C, waist circumference, and walking time were also identified as important factors for predicting MS onset among male participants aged 30 years. HDL-C and skipping breakfast were also revealed as important factors among male participants aged 35 years. For female participants aged 30 and 35 years, BMI, waist circumference, uric acid level, and triglyceride level were the most important predictors of MS onset (**Figure 3**).

**Figure 3.**
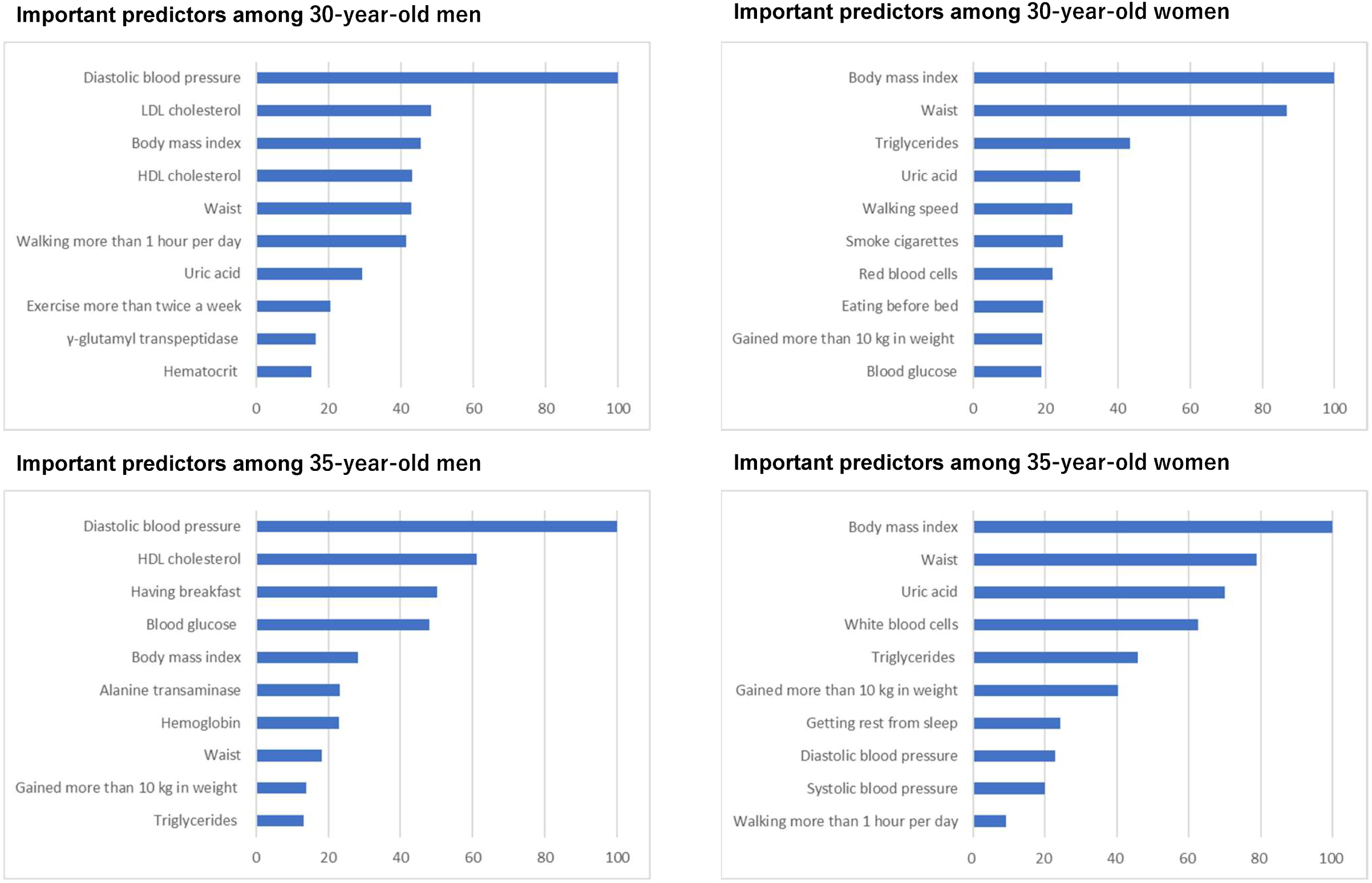
Important predictors of metabolic syndrome onset with values of important variables calculated using the random forest method.

The MDS plots of the male participants at ages 30 and 35 were divided into two major clusters, and most of the MS-onset group was included in the right cluster of the plot. On the other hand, in the MDS plots of the female participants at ages 30 and 35, non-MS cases formed a single cluster and MS-onset cases were scattered away from the non-MS-onset cluster (**Figure 4**).

**Figure 4.**
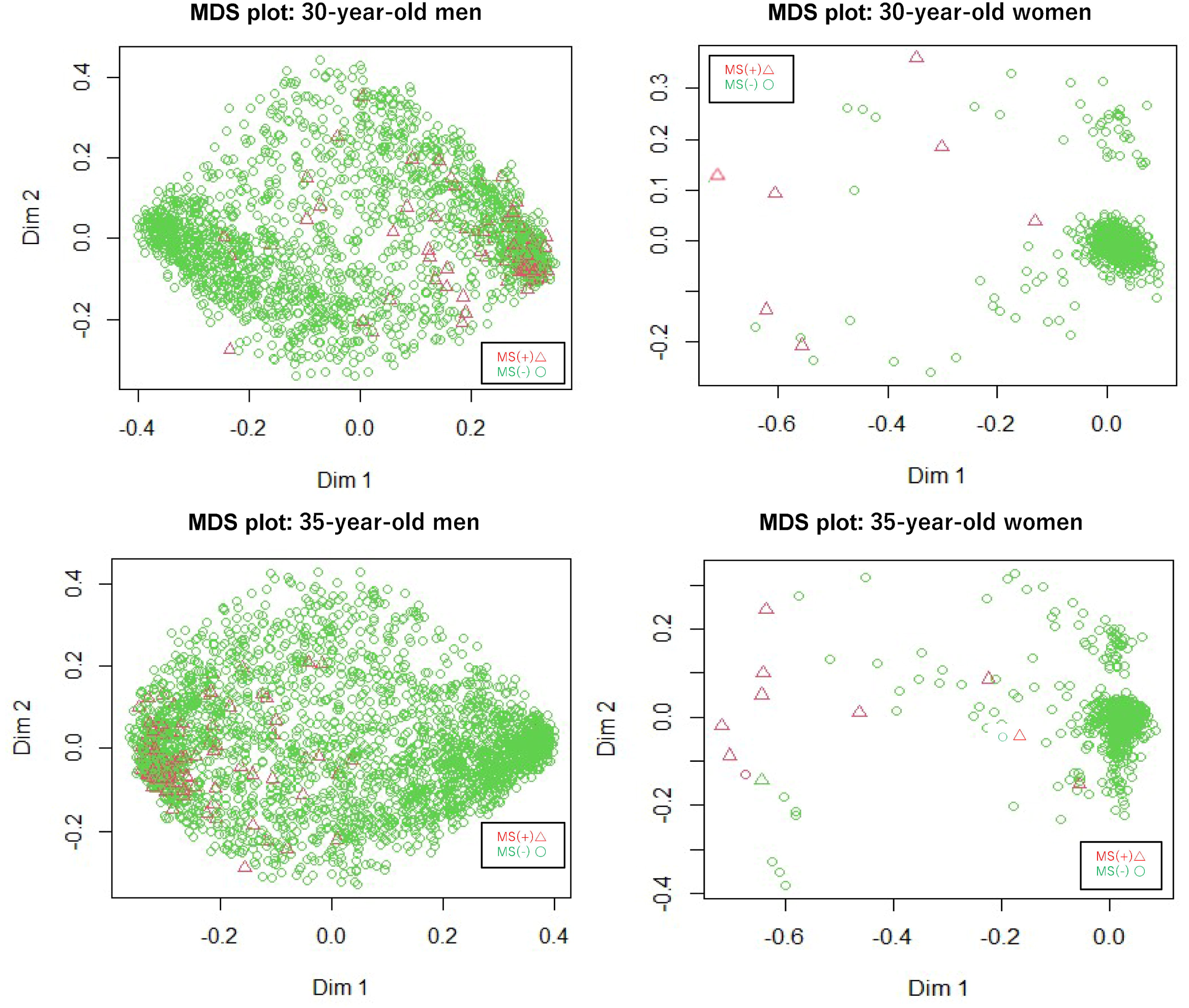
Clustering of young workers with the multidimensional scaling (MDS) plot using the random forest method.

## 4. DISCUSSION

In this study, we applied the RF and LR machine learning methods to 10-year longitudinal health checkup data from Japanese companies and developed models to predict MS onset at age 40 from health checkup data of individuals aged 30 and 35 years. The accuracy of the prediction models using RF was higher than that of models using LR. When we examined the predictors associated with MS onset, we found the most important predictors that were common to both the RF and LR models. The MDS plots using the RF method showed different characteristics between the participants with MS and those without MS onset.

In this study, the accuracy of the RF prediction model was higher than that of the LR model. When LR is used for prediction, the existence of a linear relationship is assumed between the outcome and explanatory variable. On the other hand, RF does not necessarily require this assumption because of its calculation methods.^19^ Furthermore, RF has been reported to have better performance than other methods when there are many explanatory variables and interactions between variables.^20^ In the present study, 28 explanatory variables were used to create the model, and a group of variables with interactions (e.g., BMI and waist circumference) were included. These may be why the accuracy of predicting the onset of MS was higher in the RF models than in the LR models.

In a previous study, the predictors of MS onset were investigated using regression analysis in Japanese men aged 30, 35, and 40 years, and increase in BMI was reported as the most important predictor.^10^ However, in the present study, diastolic blood pressure was presented as the most important predictor. The reason for the difference in the results may be due to multicollinearity that occurred differently in the RF and LR models. A previous study showed that RF has better performance for nonlinear relationships than regression analysis (Cox-proportional hazards regression).^21^ This characteristic of RF may lead to the presence of different factors for MS prediction.

Another previous study using the neural network method also showed diastolic blood pressure as an important predictor,^22^ and another study also reported that diastolic blood pressure is associated with the development of MS at a young age.^23^ These studies support the results of the present study. The value of diastolic blood pressure may be more useful for judging the future onset of MS than increase in BMI because blood pressure can be ascertained at one point, while increase in BMI cannot be measured at one time.

In the analyses of female participants, the most important predictor of MS was BMI, followed by abdominal circumference, as noted in previous studies.^10^Abdominal circumference is the criterion for determining the onset of MS, and the criteria for abdominal circumference are stricter in women than in men. Therefore, the importance of abdominal circumference and BMI might be higher in female participants than in male participants.

Among the questionnaire items, walking time in men at age 30, skipping breakfast in men at age 35, walking speed in women at age 30, and not feeling rested from sleep in women at age 35 were identified as important predictors. A previous study reported that daily exercise habits, regular diet, and restful sleep were associated with the development of MS in both men and women.^24^This finding is consistent with the results of the current study. To evaluate these items, we used RF with conditional inference trees to calculate the importance of variables, which enabled us to select important variables with only a few categories that were difficult to evaluate with the conventional RF.

In the MDS plot of female participants, MS cases were sporadically located away from the cluster. This sporadic population may represent a future unhealthy population. However, in the plot of male participants, MS cases were concentrated in one of two separate clusters; thus, clusters with a high concentration of MS cases may represent future unhealthy populations. The differences between MS and non-MS participants in the MDS plot were more conspicuous among female participants than among male participants. This may be because the criteria for determining MS were stricter in women than in men, and the women who will experience MS onset at age 40 had more distinctive characteristics than men in the same situation.

The strength of our study is our construction of models with machine learning methods to predict the onset of MS in men and in women using large longitudinal health examination data of young people collected over a 10-year period at a Japanese company. In addition, by using a highly interpretable machine learning method, we were able to identify important predictors from many health examination items. To our knowledge, this is the first study to develop prediction models with machine learning methods to predict the onset of MS using longitudinal data of men and women in their 30s.

### 4.1. Limitations

This study has several limitations. First, there is a possibility of selection bias because we developed and evaluated the prediction model using only the data of healthy employees in a large company. Second, it is difficult to apply the same result to populations in other countries because this study used only Japanese health examination data and the MS criteria used in Japan. Third, we could not secure a sufficient number of female subjects, and the prediction models might be less accurate for the female population than for the male population. However, we used Firth’s bias reduction method to reduce the problem of complete separation caused by the small number of cases. Fourth, the importance of some variables, especially questionnaire items, might not have been properly evaluated because the variable importance of some items with few categories tends to be underestimated in RF models. However, we used the RF method with conditional inference trees to evaluate the importance of items with few categories.

### 4.2. Clinical indication

By applying the prediction model developed in this study to the health checkup data of men and women in their 30s, it may be possible to prevent MS onset in an effective way. For instance, companies and municipalities with limited medical sources can identify high-risk groups for MS by applying our models to their data. In this study, in addition to BMI and waist circumference, diastolic blood pressure, LDL-C, and HDL-C in men and uric acid and triglyceride in women were noted as important predictors of MS. In addition, walking habits in both men and women, skipping breakfast in men, and restful sleep in women were also presented as important factors.

In this study, we could not determine a causal relationship between these factors and the development of MS. However, based on these results, we can prevent MS onset efficiently by focusing on items by sex when conducting health guidance for people in their 30s. Currently, the Japanese legal health checkup does not include items for determining MS in people in their 30s. However, by applying our prediction model to the health examination of young people, it may be possible to identify high-risk groups for MS at an early stage. Furthermore, by providing health guidance to young people at high risk, focusing on the predictors identified in this study, the onset of MS can be prevented, resulting in a reduction of the nation’s healthcare costs.

## 5. CONCLUSION

We developed a high-accuracy prediction model with a machine learning method that predicts the onset of MS at age 40 based on health examination data obtained at ages 30 and 35. Some important sex-specific predictors were identified using this highly interpretable machine learning method. Applying our models to routine healthcare management should provide early and appropriate health interventions to young people for preventing the onset of MS in this population.

## Data Availability

The data that support the findings of this study are available on request from the corresponding author. The data are not publicly available due to privacy or ethical restrictions.

## ACKNOWLEDGMENT

We are grateful to all the staff of Health Insurance Association A for preparing the dataset for the current study.

